# Evaluating the Effectiveness of 2024-2025 Seasonal mRNA-1273 Vaccination Against COVID-19-Associated Hospitalizations and Medically Attended COVID-19 among adults aged ≥ 18 years in the United States

**DOI:** 10.1101/2025.03.27.25324770

**Authors:** Amanda Wilson, Alina Bogdanov, Zhe Zheng, Taylor Ryan, Ni Zeng, Keya Joshi, Tianyi Lu, Machaon Bonafede, Andre B. Araujo

## Abstract

**Background:** This study evaluated the effectiveness of Moderna’s updated mRNA-1273 vaccine targeting the KP.2 variant, compared to people who did not receive any 2024-2025 COVID-19 vaccine, in preventing COVID-19-associated hospitalizations and medically attended COVID-19 among adults aged ≥18 years in the United States during the 2024-2025 season.

**Methods:** Data were extracted from linked administrative healthcare claims and electronic health records (EHR) for vaccinations from 23 August 2024 through 24 December 2024 and followed through 31 December 2024. We conducted a retrospective matched cohort study with propensity score weighting to adjust for differences between groups to assess vaccine effectiveness (VE) against COVID-19 outcomes. VE was calculated as 1 minus the hazard ratio (HR) from Cox proportional hazards models.

**Results:** Overall, 465,073 mRNA-1273 KP.2 vaccine recipients were matched 1:1 to unexposed adults. The mean (standard deviation) age was 63 (17) years, with more than half of the population being 65 years or older. Approximately 70% of individuals had an underlying medical condition making them high risk for severe outcomes for COVID-19. VE was 52.8% (95% confidence interval [CI], 34.8%, 65.8%) against COVID-19–related hospitalization and 39.4% (35.0%, 43.5%) against medically attended COVID-19 over a median follow-up of 57 (interquartile range: 33-78) days.

**Conclusion:** The mRNA-1273 KP.2 vaccine demonstrated significant incremental effectiveness in preventing hospitalization with COVID-19 and medically attended COVID-19 in adults during the 2024-2025 season to date. These findings support ongoing vaccination efforts to mitigate the public health impact of COVID-19.

**Funding:** This work was supported by Moderna Inc.

## Introduction

The COVID-19 pandemic has posed unprecedented challenges to global health, necessitating the development and deployment of vaccines to mitigate the public health impact of COVID-19. The initial vaccines demonstrated high efficacy in clinical trials[1,2] ; however, the emergence of variants of concern (VOCs) and increasing immunity in the population due to prior infection and vaccination have raised questions about vaccine effectiveness (VE) over time, as well as the need for vaccination targeting new VOCs. Notably, the Omicron variant and its sublineages have exhibited increased transmissibility and partial resistance to neutralization by antibodies elicited by prior infection or vaccination with the ancestral strain [3–5].

To address the evolving threat posed by SARS-CoV-2 variants, vaccine manufacturers have updated their formulations. The JN.1 family lineage emerged from BA.2.86 in late 2023 which necessitated an update from the XBB.1.5 formulation that was available beginning in August 2023. The updated vaccine aims to enhance protection against currently circulating variants, thereby reducing COVID- 19-related hospitalizations and medically attended cases. The mRNA-1273 KP.2 vaccine, developed by Moderna and available in the US as of 23 August 2024, was designed to target the Omicron KP.2 sublineage [6]. While this formulation was anticipated to cross neutralize against JN.1 family members there is limited clinical evidence to confirm this, driving the need for real-world evidence (RWE).

RWE is crucial for assessing the effectiveness of updated vaccines across diverse populations and settings. Observational studies leveraging electronic health records (EHR) and claims data provide valuable insights into VE against severe outcomes, such as hospitalization and death[7,8]. Previous RWE studies have demonstrated that updated mRNA vaccines in prior seasons offered substantial protection against severe disease caused by emerging variants[9].

This study aims to evaluate the effectiveness of the mRNA-1273 KP.2 vaccine in preventing COVID- 19-associated hospitalizations and medically attended COVID-19 among adults aged ≥18 years in the United States during the 2024–2025 season.

## Methods

### Data Source

This study leveraged electronic health record (EHR) data from the Veradigm Network EHR linked to administrative healthcare claims sourced from Komodo Health. The EHR data are sourced from ambulatory/outpatient primary care and specialty settings, and the claims data include inpatient, outpatient, and pharmacy sources. This integrated dataset has been previously characterized and used previously in COVID-19 epidemiology and VE research [10–13].

This study was conducted in accordance with the Declaration of Helsinki and relevant guidelines for observational studies. The research involved de-identified data and did not require direct patient involvement. Therefore, specific ethics approval and informed consent were not needed, as the data were anonymized and complied with patient privacy regulations. Access to the data was provided under agreements with the data provider, which ensured adherence to ethical standards, including the protection of patient confidentiality.

### Participants and Study Design

This was an observational, matched cohort study of adults in the US who were vaccinated from 23 August 2024 through 24 December 2024 (intake period) and followed through 31 December 2024. Patients vaccinated with mRNA-1273 2024-25 formulation targeting the KP.2 variant (exposed) were 1:1 matched to those without evidence of receiving a 2024-2025 COVID-19 vaccine (unexposed) on the eligible index date. The index date was defined as the date of mRNA-1273 KP.2 vaccination among the exposed and the matched date among the unexposed. To allow for establishment of vaccine-induced immunity, the cohort entry date (CED) was defined as 7 days following the index date. The pre-index period started 12 September 2023, the date the prior season’s mRNA1273 vaccine became available + 1 day. Individuals were eligible for inclusion in the study if they were ≥ 18 years at the start of the intake period and had continuous medical and pharmacy enrollment (with 45-day allowable gaps) from the start of the pre-index period through CED, regardless of prior vaccination history. Patients were excluded if they had missing or conflicting age, sex, or state of residence, evidence of a COVID-19 diagnosis or COVID-19 treatment 90 days prior to index date through CED, evidence of vaccination with any seasonal updated 2024-2025 COVID-19 vaccine from 60 days prior to index date through the end of the intake period (excluding index date, where by definition exposed patients received mRNA-1273 KP.2), receipt of any other COVID-19 vaccine from 60 days prior to index date through CED, or had <1 day of follow-up.

Subpopulations of interest were identified as age 50 years and older (at start of intake), 65 years and older (at start of intake), and individuals with underlying medical conditions associated with increased risk of severe COVID-19 outcomes, as defined by the Centers for Disease Control and Prevention (CDC)[14]. These included asthma, cancer, cerebrovascular disease, chronic kidney disease, chronic liver disease, chronic lung disease, cystic fibrosis, dementia, diabetes mellitus, disability, heart conditions, HIV, mental health conditions, obesity (body mass index > 30), physical inactivity, pregnancy, primary immunodeficiencies, respiratory tuberculosis, smoking, and solid organ or stem cell transplant, and use of select immunosuppressive medications.

### Outcomes

The primary outcome of COVID-19 hospitalization was defined as a hospitalization claim with a COVID-19 International Classification of Diseases (ICD)-10 diagnosis code (U07.1 or J12.82) in any position. The secondary outcome of medically attended COVID-19 was defined as at least one claim or record with a COVID-19 ICD-10 diagnosis code (U07.1 or J12.82) or related Systematized Nomenclature of Medicine (SNOMED) code in any position or setting (including hospital admissions as in the primary outcome, as well as emergency department visits, urgent care visits, office visits, telemedicine visits, and laboratory results). Follow-up time for the assessment of the outcomes started on CED. Patients were followed until the first occurrence of the outcome, or other censoring criteria (health plan disenrollment, vaccination with any COVID-19 vaccine during follow-up, or end of study). (Figure 1)

**Figure 1:**
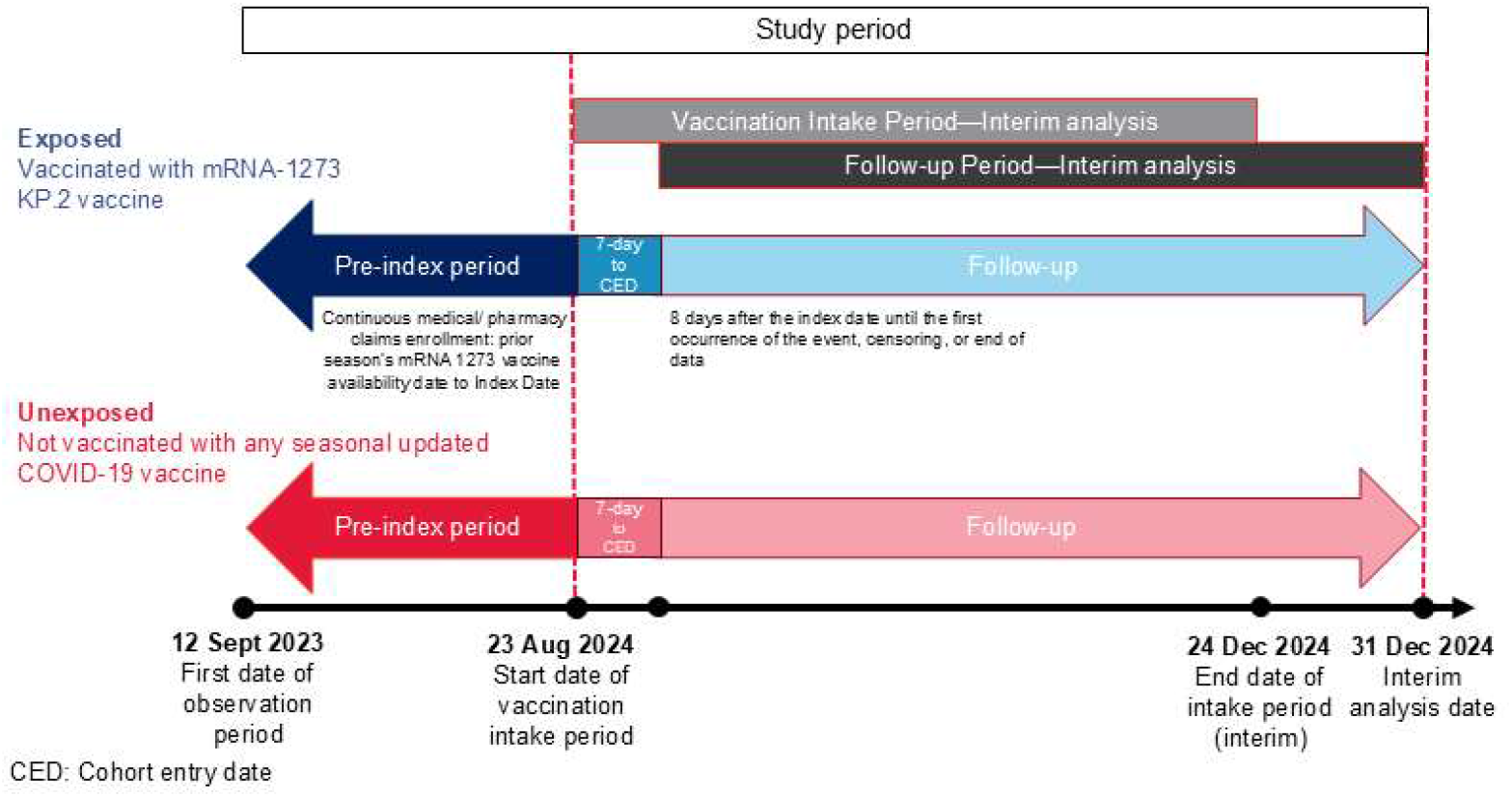
Study Design.

### Covariates

Covariates included demographic variables (age, sex, race, ethnicity, geographic region), comorbidities (e.g., diabetes, cardiovascular disease), and healthcare utilization patterns. The frequency and distribution of covariates were assessed at relevant time points prior to and/or including the index date and described between exposed and unexposed patients. Presence of high-risk and immunocompromising conditions were assessed in the pre-index period from up to 365 days prior to the index date, up through the index date (inclusive). Healthcare resource utilization was assessed from up to 365 days prior to the index date, up to one day before the index date. Influenza history and influenza vaccination history was assessed from the start of the pre-index period up to one day before the index date. COVID-19 vaccination history was assessed from the start of the pre-index period up to 60 days prior to the index date, and COVID-19 history from the start of the pre-index period up to 90 days prior to the index date. Demographic variables were assessed on the start of the intake period.

### Statistical Analysis

Unexposed individuals were 1:1 matched to the exposed based on age, sex, race, ethnicity, geographic region, week of last healthcare activity before start of intake period, evidence of vaccination with a prior season’s vaccine during the pre-index period, and the calendar date of the index date in the exposed. Matching procedures and statistical analyses were conducted for the overall cohort and each sub-population independently.

To account for potential differences in measured baseline confounders even after matching, logistic regression models were used to calculate propensity scores and corresponding inverse probability of treatment weights (IPTWs). Stabilized IPTWs were calculated as the probability of being exposed to mRNA-1273 KP.2 by the propensity score (PS) for the exposed group and as the probability of being unexposed divided by (1-PS) among the unexposed group. Sample balance before and after IPTW was assessed by calculation of the standardized mean differences (SMD). SMDs with absolute values >0.1 indicated covariate imbalance. Following IPTW, any covariates with remaining imbalance were to be included in the multivariate outcome model for additional adjustment to address potential residual confounding. Covariates were reported descriptively before and after weighting with means and standard deviations for continuous variables and number and percent for categorical variables.

Incidence rates and unadjusted HRs were reported for the unweighted sample, and adjusted HRs were reported for the weighted sample. Unadjusted HRs were calculated using a Cox regression model with mRNA-1273 KP.2 vaccine as the only predictor. Adjusted HRs were calculated for the weighted sample using a weighted Cox regression model that included mRNA-1273 KP.2 vaccination status and any covariates with an SMD greater than 0.1 after IPTW. Cumulative incidence plots with 95% confidence intervals were generated to visually assess the vaccine effectiveness changes over time to assess the proportional hazards assumption. Subsequently, the unadjusted and adjusted VE for each outcome were calculated as (1-HR) * 100% and reported with 95% CIs.

Data cleaning and analytic file generation was conducted via SQL. Statistical analyses were conducted using SAS V9.4.

## Results

Overall, 465,073 mRNA-1273 KP.2 recipients were matched 1:1 to unexposed adults (Figure 2). The exposed and unexposed cohorts were well balanced after weighting, with no SMDs greater than 0.1.

**Figure 2.**
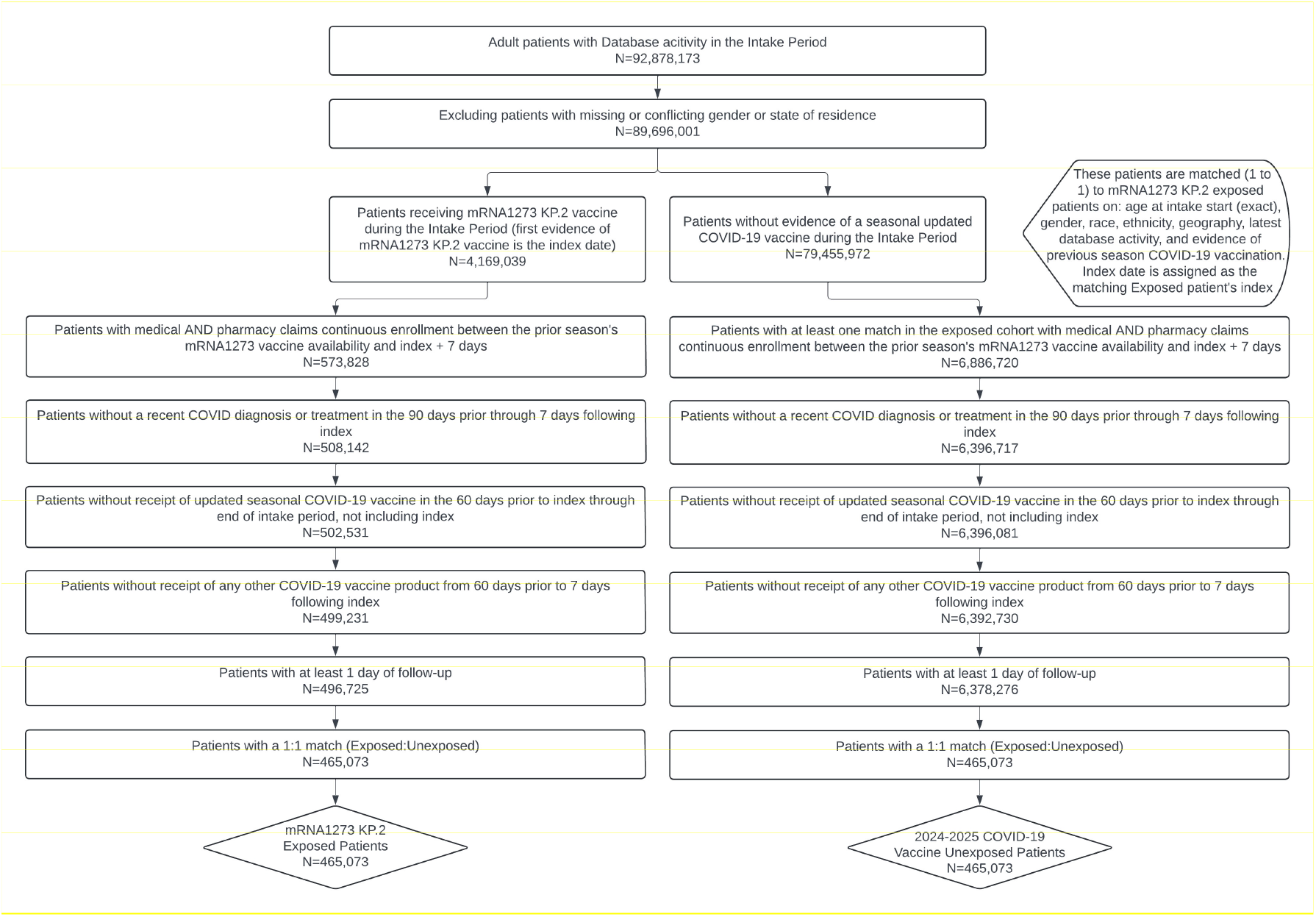
Study Population.

The mean (standard deviation) age in both cohorts was 63 (17) years; 57.8% were female. 78.7% of the study population was at least 50 years old and 55.6% were at least 65 years old. Most patients (73%) in the study population had documentation of a previous vaccination for COVID-19 from the start of the pre-index period up to 60 days prior to index. Over 87% had their most recent documented activity with the healthcare system within 4 weeks prior to the index date. Most vaccinations during the study period occurred during September (41.7%) or October (38.0%). Pre-weighting, approximately 70% had at least 1 underlying medical condition associated with an increased risk of severe COVID-19 outcomes. The most common underlying medical conditions pre-weighting were obesity (exposed: 25.3% and unexposed: 26.5%), diabetes mellitus (21.7% and 23.5%, respectively), and heart conditions (17.9% and 19.7%, respectively). See Table 1.

**Table 1.** Baseline Characteristics (pre-weighting and post-weighting, and SMDs)

|  | Pre-weighting |  |  | Post-weighting |  |  |
| --- | --- | --- | --- | --- | --- | --- |
|  | Exposed | Unexposed | SMD | Exposed | Unexposed | SMD |
| **Number of Patients** | 465,073 | 465,073 |  | 463,093 | 464,169 |  |
| **Age; mean (SD)** | 63 (17.0) | 63 (17.0) | 0.0000 | 63 (16.9) | 63 (16.9) | 0.0004 |
| **Age Categories; n (%)** |  |  | 0.0000 |  |  | 0.0014 |
| 18-24 | 10,592 (2.3) | 10,592 (2.3) |  | 10,456 (2.3) | 10,479 (2.3) |  |
| 25-29 | 11,377 (2.4) | 11,377 (2.4) |  | 11,245 (2.4) | 11,269 (2.4) |  |
| 30-34 | 16,672 (3.6) | 16,672 (3.6) |  | 16,485 (3.6) | 16,518 (3.6) |  |
| 35-39 | 19,822 (4.3) | 19,822 (4.3) |  | 19,637 (4.2) | 19,663 (4.2) |  |
| 40-44 | 20,637 (4.4) | 20,637 (4.4) |  | 20,474 (4.4) | 20,494 (4.4) |  |
| 45-49 | 20,489 (4.4) | 20,489 (4.4) |  | 20,338 (4.4) | 20,351 (4.4) |  |
| 50-54 | 24,939 (5.4) | 24,939 (5.4) |  | 24,775 (5.3) | 24,788 (5.3) |  |
| 55-59 | 33,093 (7.1) | 33,093 (7.1) |  | 32,884 (7.1) | 32,904 (7.1) |  |
| 60-64 | 49,354 (10.6) | 49,354 (10.6) |  | 49,126 (10.6) | 49,149 (10.6) |  |
| 65-69 | 52,961 (11.4) | 52,961 (11.4) |  | 52,803 (11.4) | 52,972 (11.4) |  |
| 70-74 | 69,858 (15.0) | 69,858 (15.0) |  | 69,591 (15.0) | 69,892 (15.1) |  |
| 75-79 | 62,435 (13.4) | 62,435 (13.4) |  | 62,357 (13.5) | 62,592 (13.5) |  |
| 80-84 | 39,934 (8.6) | 39,934 (8.6) |  | 39,932 (8.6) | 40,058 (8.6) |  |
| 85-89 | 32,910 (7.1) | 32,910 (7.1) |  | 32,990 (7.1) | 33,042 (7.1) |  |
| **Sex; n (%)** |  |  |  |  |  |  |
| Male | 196,255 (42.2) | 196,255 (42.2) | 0.0000 | 195,438 (42.2) | 195,869 (42.2) | 0.0001 |
| **Race; n (%)** |  |  | 0.0000 |  |  | 0.0006 |
| Black | 24,812 (5.3) | 24,812 (5.3) |  | 24,756 (5.3) | 24,769 (5.3) |  |
| Other | 67,999 (14.6) | 67,999 (14.6) |  | 67,794 (14.6) | 67,908 (14.6) |  |
| White | 281,593 (60.5) | 281,593 (60.5) |  | 280,455 (60.6) | 281,206 (60.6) |  |
| Unknown | 90,669 (19.5) | 90,669 (19.5) |  | 90,088 (19.5) | 90,286 (19.5) |  |
| **Ethnicity; n (%)** |  |  | 0.0000 |  |  | 0.0000 |
| Hispanic | 8,503 (1.8) | 8,503 (1.8) |  | 8,477 (1.8) | 8,476 (1.8) |  |
| Non-Hispanic | 282,192 (60.7) | 282,192 (60.7) |  | 281,048 (60.7) | 281,719 (60.7) |  |
| Unknown | 174,378 (37.5) | 174,378 (37.5) |  | 173,568 (37.5) | 173,974 (37.5) |  |
| **Insurance Type; n (%)** |  |  | 0.1633 |  |  | 0.0067 |
| Commercial | 209,648 (45.1) | 200,904 (43.2) |  | 203,883 (44.0) | 203,915 (43.9) |  |
| Medicaid | 11,656 (2.5) | 16,883 (3.6) |  | 14,024 (3.0) | 14,167 (3.1) |  |
| Medicare Advantage | 223,474 (48.1) | 210,871 (45.3) |  | 217,339 (46.9) | 217,654 (46.9) |  |
| Medicare FFS | 18,698 (4.0) | 32,799 (7.1) |  | 25,398 (5.5) | 25,830 (5.6) |  |
| Unknown | 1,597 (0.3) | 3,616 (0.8) |  | 2,451 (0.5) | 2,604 (0.6) |  |
| **Region; n (%)** |  |  | 0.0000 |  |  | 0.0007 |
| Midwest | 77,810 (16.7) | 77,810 (16.7) |  | 77,509 (16.7) | 77,603 (16.7) |  |
| Northeast | 157,593 (33.9) | 157,593 (33.9) |  | 156,808 (33.9) | 157,212 (33.9) |  |
|  | Exposed | Unexposed | SMD | Exposed | Unexposed | SMD |
| South | 121,577 (26.1) | 121,577 (26.1) |  | 121,123 (26.2) | 121,394 (26.2) |  |
| West | 108,090 (23.2) | 108,090 (23.2) |  | 107,650 (23.2) | 107,957 (23.3) |  |
| Unknown | 3 (0.0) | 3 (0.0) |  | 3 (0.0) | 3 (0.0) |  |
| **Month of Index Date; n (%)** |  |  | 0.0000 |  |  | 0.0236 |
| 2024-08 | 19,314 (4.2) | 19,314 (4.2) |  | 19,896 (4.3) | 19,062 (4.1) |  |
| 2024-09 | 194,002 (41.7) | 194,002 (41.7) |  | 196,016 (42.3) | 192,455 (41.5) |  |
| 2024-10 | 176,564 (38.0) | 176,564 (38.0) |  | 173,872 (37.5) | 176,489 (38.0) |  |
| 2024-11 | 63,891 (13.7) | 63,891 (13.7) |  | 62,376 (13.5) | 64,549 (13.9) |  |
| 2024-12 | 11,302 (2.4) | 11,302 (2.4) |  | 10,933 (2.4) | 11,614 (2.5) |  |
| **Number of OP visits; mean (SD)** | 2.4 (6.0) | 2.6 (6.5) | 0.0337 | 2.5 (6.3) | 2.5 (6.2) | 0.0009 |
| **Number of hospitalizations; mean (SD)** | 0.1 (0.4) | 0.1 (0.5) | 0.0709 | 0.1 (0.5) | 0.1 (0.5) | 0.0028 |
| **Baseline Clinical Comorbidities; n (%)** |  |  |  |  |  |  |
| Cancer | 48,432 (10.4) | 49,556 (10.7) | 0.0079 | 48,896 (10.6) | 49,059 (10.6) | 0.0003 |
| Cerebrovascular disease | 32,946 (7.1) | 37,837 (8.1) | 0.0397 | 35,307 (7.6) | 35,513 (7.7) | 0.0010 |
| Chronic kidney disease | 56,802 (12.2) | 61,331 (13.2) | 0.0292 | 59,100 (12.8) | 59,324 (12.8) | 0.0006 |
| Chronic lung diseases | 40,497 (8.7) | 46,365 (10.0) | 0.0434 | 43,369 (9.4) | 43,592 (9.4) | 0.0009 |
| Chronic liver diseases | 5,542 (1.2) | 6,201 (1.3) | 0.0127 | 5,844 (1.3) | 5,876 (1.3) | 0.0004 |
| Cystic fibrosis | 84 (0.0) | 82 (0.0) | 0.0003 | 84 (0.0) | 84 (0.0) | 0.0000 |
| Diabetes mellitus, type 1 and type 2 | 101,134 (21.7) | 109,396 (23.5) | 0.0425 | 105,225 (22.7) | 105,545 (22.7) | 0.0004 |
| Disabilities | 34,575 (7.4) | 33,952 (7.3) | 0.0051 | 34,210 (7.4) | 34,258 (7.4) | 0.0003 |
| Heart conditions | 83,216 (17.9) | 91,481 (19.7) | 0.0455 | 87,159 (18.8) | 87,567 (18.9) | 0.0011 |
| HIV | 2,179 (0.5) | 1,803 (0.4) | 0.0124 | 1,999 (0.4) | 1,989 (0.4) | 0.0005 |
| Mental health disorders | 77,216 (16.6) | 81,179 (17.5) | 0.0227 | 79,081 (17.1) | 79,333 (17.1) | 0.0004 |
| Neurologic conditions | 13,795 (3.0) | 18,862 (4.1) | 0.0592 | 16,197 (3.5) | 16,373 (3.5) | 0.0016 |
| Obesity | 117,729 (25.3) | 123,358 (26.5) | 0.0276 | 120,206 (26.0) | 120,500 (26.0) | 0.0001 |
| Primary Immunodeficiencies | 8,380 (1.8) | 8,771 (1.9) | 0.0062 | 8,583 (1.9) | 8,608 (1.9) | 0.0001 |
| Pregnancy and recent pregnancy | 1,344 (0.3) | 1,227 (0.3) | 0.0048 | 1,261 (0.3) | 1,263 (0.3) | 0.0000 |
| Physical inactivity | 445 (0.1) | 475 (0.1) | 0.0021 | 455 (0.1) | 458 (0.1) | 0.0001 |
| Smoking, current and former | 64,145 (13.8) | 72,443 (15.6) | 0.0504 | 67,975 (14.7) | 68,322 (14.7) | 0.0012 |
|  | Exposed | Unexposed | SMD | Exposed | Unexposed | SMD |
| Solid organ or hematopoietic cell transplantation | 1,302 (0.3) | 1,410 (0.3) | 0.0043 | 1,357 (0.3) | 1,364 (0.3) | 0.0001 |
| Tuberculosis | 131 (0.0) | 144 (0.0) | 0.0016 | 139 (0.0) | 138 (0.0) | 0.0002 |
| Use of immunosuppressive medications | 28,922 (6.2) | 26,706 (5.7) | 0.0201 | 27,820 (6.0) | 27,842 (6.0) | 0.0004 |
| Asthma | 41,930 (9.0) | 40,502 (8.7) | 0.0108 | 41,149 (8.9) | 41,214 (8.9) | 0.0002 |
| **Time since last COVID monovalent vaccination; n (%)** |  |  | 0.1411 |  |  | 0.0451 |
| <= 90 days | 1,439 (0.3) | 7,408 (1.6) |  | 2,597 (0.6) | 4,428 (1.0) |  |
| 91-180 day | 18,230 (3.9) | 22,355 (4.8) |  | 20,266 (4.4) | 20,326 (4.4) |  |
| 180+ days | 322,112 (69.3) | 311,618 (67.0) |  | 318,214 (68.7) | 317,933 (68.5) |  |
| Not reported | 123,292 (26.5) | 123,692 (26.6) |  | 122,017 (26.3) | 121,482 (26.2) |  |
| **Time since last COVID infection; n (%)** |  |  | 0.0454 |  |  | 0.0021 |
| <= 120 days | 2,524 (0.5) | 2,319 (0.5) |  | 2,406 (0.5) | 2,405 (0.5) |  |
| 121-180 day | 2,551 (0.5) | 2,141 (0.5) |  | 2,335 (0.5) | 2,328 (0.5) |  |
| 180+ days | 19,086 (4.1) | 15,323 (3.3) |  | 17,111 (3.7) | 16,936 (3.6) |  |
| Not reported | 440,912 (94.8) | 445,290 (95.7) |  | 441,242 (95.3) | 442,500 (95.3) |  |
| **Evidence of COVID vaccination with prior season's vaccine during pre-index period; n (%)** | 340,775 (73.3) | 340,775 (73.3) | 0.0000 | 340,259 (73.5) | 341,964 (73.7) | 0.0045 |
| **Evidence of influenza vax from the start of the pre-index period to 1 day before index; n (%)** | 356,669 (76.7) | 306,347 (65.9) | 0.2409 | 329,378 (71.1) | 329,948 (71.1) | 0.0009 |
| **Diagnosis of influenza from start of pre-index period until one day before index; n (%)** | 6,966 (1.5) | 7,258 (1.6) | 0.0051 | 7,077 (1.5) | 7,098 (1.5) | 0.0001 |
| **Week of last healthcare activity before start of intake; n (%)** |  |  | 0.0000 |  |  | 0.0011 |
| 0-4 weeks | 408,052 (87.7) | 408,052 (87.7) |  | 406,626 (87.8) | 407,674 (87.8) |  |
| 5-26 weeks | 52,126 (11.2) | 52,126 (11.2) |  | 51,636 (11.2) | 51,686 (11.1) |  |
| 27-52 weeks | 4,074 (0.9) | 4,074 (0.9) |  | 4,009 (0.9) | 3,991 (0.9) |  |
| > 52 weeks or unknown | 821 (0.2) | 821 (0.2) |  | 823 (0.2) | 818 (0.2) |  |

The overall median (interquartile range) follow-up time was 55 (32-77) days. A total of 51 COVID-19– related hospitalizations and 1,270 COVID-19–related medical encounters were identified pre- weighing in the exposed, and 118 and 2,016 in the unexposed, respectively. After adjustment, the incidence of COVID-19 hospitalizations was 0.01% in the exposed cohort and 0.02% in the unexposed cohort over the follow-up period. The incidence of medically attended COVID-19 was 0.27% in the exposed cohort and 0.44% in the unexposed cohort over the follow-up period.

Cumulative incidence curves for hospitalizations and medically-attended COVID-19 are presented in Figure 3.

**Figure 3.**
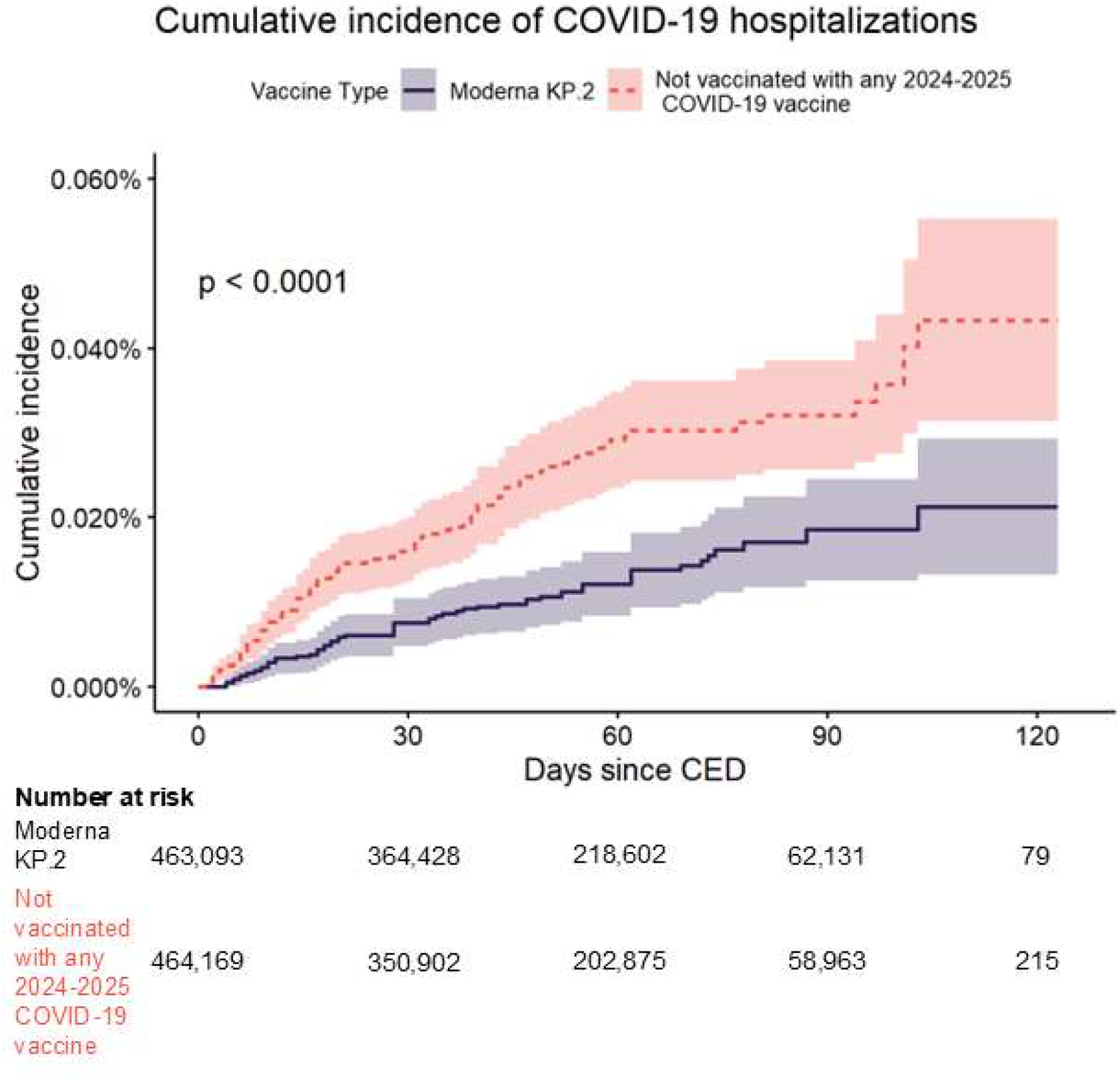
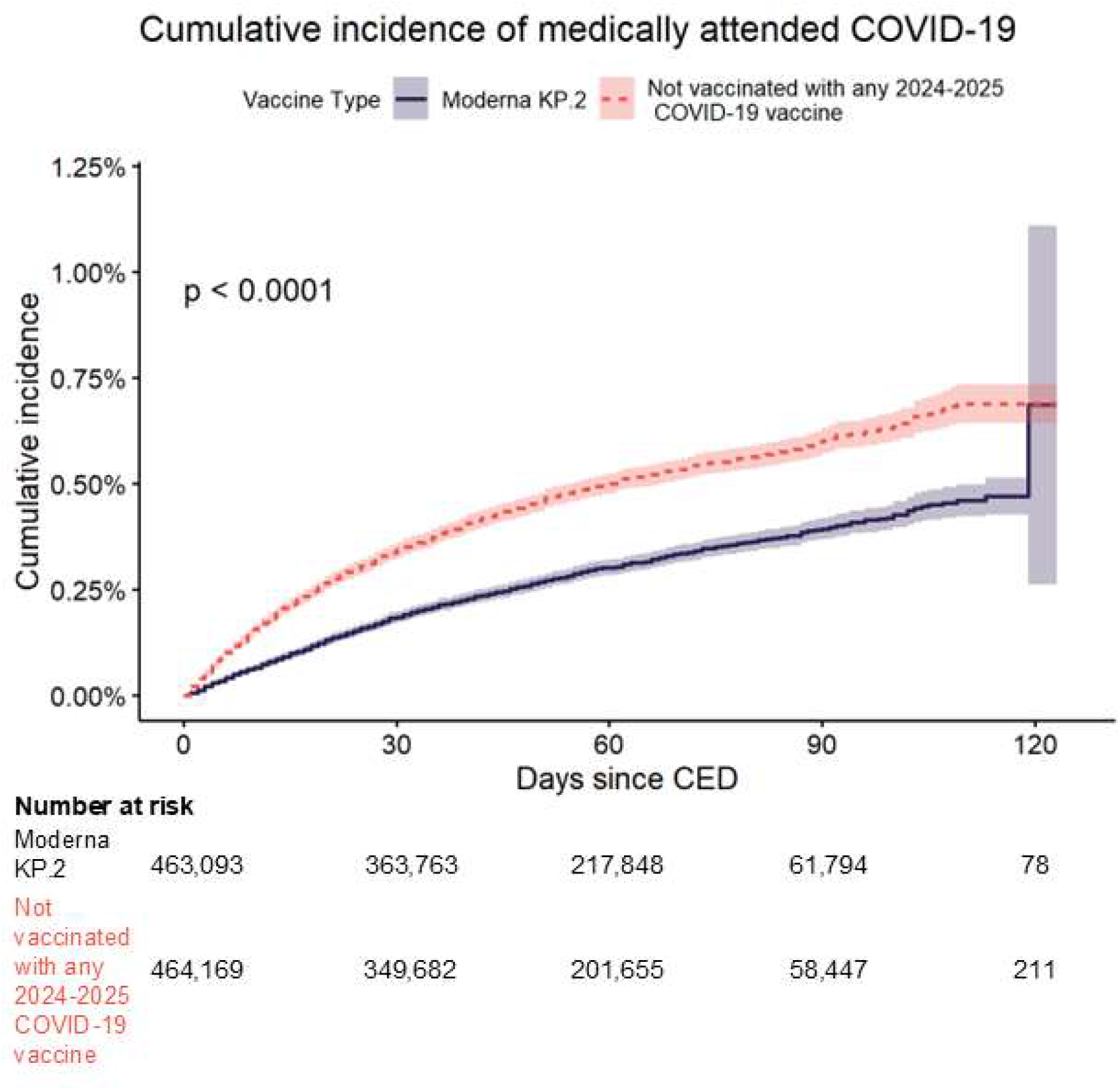
Cumulative incidence curves of time to 1) COVID-19 -Related Hospitalizations, and 2) Medically attended COVID.

Adjusted overall VE (95% CI) was 52.8% (34.8%, 65.8%) against COVID-19–related hospitalization and 39.4% (35.0%, 43.5%) against medically attended COVID-19. Adjusted VE (95% CI) against COVID-19–related hospitalization was 53.1% (35.1%, 66.2%) for adults ≥50 years old, 53.2% (34.2%, 66.7%) among adults ≥65 years old, and 46.5% (25.3%, 61.7%) among adults with at least 1 underlying medical condition. Adjusted VE (95% CI) against medically attended COVID-19 was 41.2% (36.5%, 45.4%) for adults ≥50 years old, 46.7% (41.7%, 51.2%) among adults ≥65 years old, and 40.4% (35.5%, 44.9%) among adults with at least 1 underlying medical condition (Table 2).

**Table 2.** Incidence Rates and Vaccine Effectiveness of 1) COVID-19 -Related Hospitalizations, and 2) Medically-attended COVID-19.

|  | Exposed Incidence; n (%) | Unexposed Incidence; n (%) | Unadjusted HR | Adjusted HR | Adjusted VE |
| --- | --- | --- | --- | --- | --- |
| **18+** |  |  |  |  |  |
| COVID-19 Hospitalization | 51 (0.01%) | 118 (0.03%) | 0.42 (0.30, 0.58) | 0.47 (0.34, 0.65) | 52.8% (34.8%, 65.8%) |
| Medically-attended COVID-19 | 1,270 (0.27%) | 2,016 (0.43%) | 0.61 (0.57, 0.65) | 0.61 (0.57, 0.65) | 39.4% (35.0%, 43.5%) |
| **50+** |  |  |  |  |  |
| COVID-19 Hospitalization | 50 (0.01%) | 116 (0.03%) | 0.41 (0.30, 0.58) | 0.47 (0.34, 0.65) | 53.1% (35.1%, 66.2%) |
| Medically-attended COVID-19 | 1,080 (0.30%) | 1,771 (0.48%) | 0.59 (0.54, 0.63) | 0.59 (0.55, 0.64) | 41.2% (36.5%, 45.4%) |
| **65+** |  |  |  |  |  |
| COVID-19 Hospitalization | 46 (0.02%) | 106 (0.04%) | 0.42 (0.30, 0.59) | 0.47 (0.33, 0.66) | 53.2% (34.2%, 66.7%) |
| Medically-attended COVID-19 | 737 (0.29%) | 1,346 (0.52%) | 0.53 (0.48, 0.58) | 0.53 (0.49, 0.58) | 46.7% (41.7%, 51.2%) |
| **18+ High Risk** |  |  |  |  |  |
| COVID-19 Hospitalization | 50 (0.02%) | 102 (0.03%) | 0.48 (0.34, 0.68) | 0.54 (0.38, 0.75) | 46.5% (25.3%, 61.7%) |
| Medically-attended COVID-19 | 992 (0.30%) | 1,643 (0.49%) | 0.59 (0.55, 0.64) | 0.60 (0.55, 0.65) | 40.4% (35.5%, 44.9%) |

## Discussion

This study’s findings align with previous evaluations of prior-years’ updated mRNA COVID-19 vaccines against hospitalization with COVID-19, reinforcing the continued importance of ongoing COVID-19 vaccinations. Protection was consistently observed across subgroups, including older adults and those with other specific underlying medical conditions, such as immunocompromised conditions, that may increase the risk of severe COVID-19. These findings of an overall VE of 52.8%, in addition to other published estimates of VE against hospitalization ranging from 45% (95%

CI 36%–53%) to 46% (95% CI 26%–60%) in immunocompetent adults aged 65 and older in the Center for Disease Control’s VISION and IVY networks [15], and 68% (95% CI 42%–82%) in a test-negative case-control study within the US Veterans Affairs Healthcare System[16], support the need for recommendations for COVID-19 vaccination with the most updated vaccine, These VE estimates underscore the benefit of administering mRNA-1273 KP.2 in the adult population and among those who are at higher risk for COVID-19-related outcomes.

By December 2024, XEC, a hybrid of two JN.1 variants, had become the predominant variant in the United States, co-circulating with KP.3.1.1, while LP.8.1, a descendant of KP.1.1.3, was on the rise. The 2024-25 vaccine is expected to be effective against the currently predominant variants (such as XEC and KP.3.1.1) and is also anticipated to work well against currently increasing variants that are likely to dominate in the future, such as LP.8.1[17]. Since the start of the COVID-19 pandemic, most of the adult population has developed immune responses against the SARS-CoV-2 virus, whether through previous infections, vaccinations, or a combination of both.[18] The current study included individuals regardless of their vaccination and infection history, which were included as matching factors or covariates. Most individuals in the study (over 70%) previously received a 2023-2024 mRNA-1273 vaccine targeting the XBB.1.5 variant. Thus, the results of this study should be interpreted in the context of the incremental protection against hospitalization provided by the most updated COVID-19 against the circulating variants in a real-world setting regardless of exposure and/or vaccination history. In an environment of low COVID-19 vaccination coverage rates across all ages, it is essential to communicate the additive protection to increase vaccine confidence among clinicians and the general population.

### Strengths

This retrospective, observational cohort study implemented multiple analytic approaches to adjust for confounding, specifically the underlying differences between the unexposed cohort and the cohort who received a vaccination with mRNA-1273 KP.2 in the Fall of 2024. Direct matching on clinical and demographic criteria such as age, sex, race, geographic region, and 2023-2024 XBB.1.5 vaccine receipt ensured balance on key confounders. Additionally, individuals were matched on week of last healthcare activity prior to the start of the intake period to mitigate potential surveillance bias by ensuring the groups are similar in terms of how often they engage with the healthcare system. The use of IPTW was able to further mitigate confounding and estimate average treatment effect among the full population; no further adjustment was needed to balance the cohorts.

A matched cohort study offers several distinct advantages over other study designs. First, it limits selection bias by identifying individuals before they experience the outcomes of interest, minimizing the impact of unmeasured confounders related to healthcare access and differential testing patterns. Finally, cohort studies allow for stronger inferences by allowing the estimation of incidence rates, which offer crucial context for VE estimates. For instance, the relatively low incidence of COVID-19 hospitalizations in this analysis compared to 2023-24 [10,19] may contribute to the lower VE estimates observed here. Furthermore, by establishing temporality, cohort studies also strengthen causal inferences and allow for the assessment of VE over time.

### Limitations

Although we have identified and adjusted for many baseline confounders, due to the lack of randomization, there could be potential residual confounding. For instance, it’s possible that exposed people had otherwise healthier lifestyles not captured in the database (i.e. beyond what is captured with healthcare utilization and measures of comorbidity), contributing to their lower rates of hospitalization with COVID-19. This is an inherent limitation in any observational study design.

The study population relies on individuals engaged with the healthcare system, possibly excluding others due to barriers such as trust, socioeconomic status, or accessibility. The outcome definition of hospitalization with COVID-19 was chosen to partially address these limitations, where a severe outcome would be less subjective and less influenced by these factors.

As in all claims and/or EHR datasets, vaccination is likely under captured; this may have led to a bias toward the null, resulting in the estimated VE being lower than true VE. Potential bias due to misclassification of the outcome and exposure is possible—diagnostic and procedural codes used to define vaccination and COVID-19-related hospitalizations may be inaccurately recorded, leading to incorrect classification of patient conditions or treatments. Additionally, observational data may not capture all hospitalizations, particularly among specific subpopulations such as uninsured individuals, underrepresented groups, or those receiving care outside the insurance network.

However, a validation study by Kadri et al.[20] reported a high positive predictive value (PPV) of 98% for the U07.1 code, used for COVID-19 diagnoses, based on clinical documentation and laboratory results.

## Conclusion

The updated 2024-2025 mRNA-1273 KP.2 vaccine demonstrated substantial protection against hospitalization and medically-attended COVID-19 during the initial months of vaccination availability in the US. These findings support continued vaccination efforts to reduce the burden of COVID-19, particularly among high-risk populations. These data support programs to improve COVID-19 vaccination coverage.

## Data Availability

Individual-level data reported in this study are not publicly shared. Upon request, and subject to review, Veradigm may provide the deidentified aggregate-level data that support the findings of this study. Deidentified data (including participant data as applicable) may be shared upon approval of an analysis proposal and a signed data access agreement. Individual-level data reported in this study are shared fully with regulatory agencies.

## Ethical Approval

This study was conducted in accordance with the Declaration of Helsinki and relevant guidelines for observational studies. The research involved de-identified claims data and did not require direct patient involvement. Therefore, specific ethics approval and informed consent were not needed, as the data were anonymized and complied with patient privacy regulations. Access to the data was provided under agreements with the data provider, which ensured adherence to ethical standards, including the protection of patient confidentiality.

## Medical Writing/Editorial Assistance

The authors acknowledge that artificial intelligence was used in the drafting of the manuscript. All text was checked and revised by authors.

