## Supplemental Tables for "Evaluating the Effectiveness of 2024-2025 Seasonal mRNA-1273 Vaccination Against COVID-19-Associated Hospitalizations and Medically Attended COVID-19 among adults aged ≥ 18 years in the United States"

### Supplementary Tables and Figures

#### Supplementary Table 1. List of codes used to identify 2024-2025 KP.2/JN.1 COVID-19 vaccines from the Veradigm EHR and linked claims datasets.

| COVID-19 vaccine type | CPT | CVX | NDC |
|---|---|---|---|
| mRNA-1273 KP.2 vaccine | 91321, 91322 | 311, 312 | 80777-0110-95, 80777-0110-96, 80777-0110-93, 80777-0110-04, 80777-0110-01, 80777-0291-80, 80777-0291-09, 80777-0291-81 |
| BNT162b2 KP.2 vaccine | 91318, 91319, 91320 | 308, 309, 310 | 00069-2432-10, 00069-2432-01, 00069-2403-10, 00069-2403-01, 59267-4438-02, 59267-4438-01, 59267-4426-02, 59267-4426-01 |
| 2024-2025 Novavax updated vaccine | 91304 | 313 | 80631-0107-01, 80631-0107-10 |

CPT, Current Procedural Terminology; CVX, vaccine administered codes; NDC, National Drug Codes

#### Supplementary Table 2. List of CVX, CPT, and NDC codes used to identify COVID-19 vaccines other than 2024-2025 updated KP.2/JN.1 formulations from the Veradigm EHR and linked claims datasets.

| Manufacturer | CPT | CVX | NDC |
|---|---|---|---|
| AstraZeneca | 91302 | 210 | 00310-1222-10, 00310-1222-15 |
| Janssen | 91303 | 212 | 59676-0580-05, 59676-0580-15 |
| Moderna | 91301, 91306, 91309, 91311, 91313, 91314, 91316, | 207, 221, 227, 228, 229, 230, 311, 312, 519 | 61434-0043-02, 80777-0100-11, 80777-0100-15, 80777-0100-98, 80777-0100-99, 80777-0102-01, 80777-0102-04, 80777-0102-93, 80777-0102-95, 80777-0102-96, 80777-0273-10, 80777-0273-15, 80777-0273-98, 80777-0273-99, 80777-0275-05, 80777-0275-99, 80777-0277-05, 80777-0277-99, 80777-0279-05, 80777-0279-99, 80777-0280-05, |

| | | | |
|---|---|---|---|
| | 91321, 91322 | | 80777-0280-99, 80777-0282-05, 80777-0282-99, 80777-0283-02, 80777-0283-99, 80777-0287-07, 80777-0287-92 |
| Novavax | 91304 | 211, 313 | 80631-0100-01, 80631-0100-10, 80631-0102-01, 80631-0102-10, 80631-0105-01, 80631-0105-02, 80631-1000-01 |
| Pfizer | 91300, 91305, 91307, 91308, 91312, 91315, 91317, 91318, 91319, 91320 | 208, 217, 218, 219, 300, 301, 302, 308, 309, 310, 520 | 00069-1000-01, 00069-1000-02, 00069-1000-03, 00069-2025-01, 00069-2025-10, 00069-2025-25, 00069-2362-01, 00069-2362-10, 00069-2377-01, 00069-2377-01, 00069-2377-10, 00069-2377-10, 00069-2392-01, 00069-2392-10, 59267-0078-01, 59267-0078-02, 59267-0078-04, 59267-0304-01, 59267-0304-02, 59267-0565-01, 59267-0565-02, 59267-0609-01, 59267-0609-02, 59267-1000-01, 59267-1000-02, 59267-1000-03, 59267-1025-01, 59267-1025-02, 59267-1025-03, 59267-1025-04, 59267-1055-01, 59267-1055-02, 59267-1055-04, 59267-1404-01, 59267-1404-02, 59267-4315-01, 59267-4315-02, 59267-4331-01, 59267-4331-02 |
| Sanofi | 91310 | 225, 226 | 49281-0618-20, 49281-0618-78 |
| Not specified | | 213 | |

CPT, Current Procedural Terminology; CVX, vaccine administered codes; ICD-10-PCS, International Classification of Disease, 10th edition, Procedure Coding System; NDC, National Drug Codes

**Supplementary Table 3. Codes used to identify COVID-19 diagnosis or treatment**

| Category | Code Type | Codes for COVID-related medical encounters |
|---|---|---|
| COVID-19 treatment | Generic drug name | abatacept, anakinra, bamlanivimab, baricitinib, bebtelovimab, betamethasone, budesonide, bupivacaine/dexamethasone, casirivimab/imdevimab, cortisone acetate, deflazacort, dexamethasone, hydrocortisone, infliximab, methylprednisolone, molnupiravir, nirmatrelvir/ritonavir, prednisolone, prednisone, remdesivir, sotrovimab, tocilizumab, triamcinolone, vilobelimab |
| COVID-19 treatment | HCPCS | M0245, M0246, Q0245, M0222, M0223, Q0222, M0240, M0241, M0243, M0244, Q0240, Q0243, Q0244, J0248, M0247, M0248, Q0247, J3262, M0249, M0250, Q0249, J0129, J1745, Q5102, Q5103, Q5104, Q5109, Q5121, C9469, J0702, J1020, J1030, J1040, J1094, J1100, J1700, J1710, J1720, J2650, J2920, J2930, J3300, J3301, J3302, J3303, J3304, J7509, J7510, J8540, Q9993 |
| COVID-19 treatment | ICD-10-PCS | XW033F6, XW043F6, XW033E5, XW043E5, XW033H5, XW043H5 |
| COVID-19 diagnosis | ICD-10-CM | J1282, U071 |

| | | |
|---|---|---|
| COVID-19 diagnosis | SNOMED | 1119302008, 119731000146105, 119741000146102, 119751000146104, 119981000146107, 1240411000000107, 1240521000000100, 1240531000000103, 1240541000000107, 1240561000000108, 1240581000000104, 674814021000119106, 840533007, 840534001, 840536004, 840539006, 866151004, 866152006, 870588003, 870589006, 870590002, 870591003, 871562009 |

CPT, Current Procedural Terminology; HCPCS, Healthcare Common Procedure Coding System; ICD-10-PCS, International Classification of Disease, 10th edition, Procedure Coding System; ICD-10-CM, International Classification of Disease, 10th edition, Clinical Modification, SNOMED, Systemized Nomenclature of Medicine
